# Spatio-Temporal Landscape of Whole-Genome DNA Methylation Patterns in Ovarian Cancer

**DOI:** 10.64898/2026.03.22.26348877

**Authors:** Giovanni Marchi, Kari Lavikka, Yilin Li, Veli-Matti Isoviita, Giulia Micoli, Daria Afenteva, Elsi Pöllänen, Susanna Holmström, Antti Häkkinen, Elina Valkonen, Ilari Maarala, Jaakko Astrén, Chiara Facciotto, Felix Dietlein, Sakari Hietanen, Taru A. Muranen, Jaana Oikkonen, Johanna Hynninen, Anni Virtanen, Alexandra Lahtinen, Sampsa Hautaniemi

## Abstract

Understanding how epigenetic mechanisms shape metastatic dissemination and therapy response in ovarian high-grade serous carcinoma (HGSC) has been hindered by the absence of large, anatomically diverse, and longitudinal datasets. Here, we present a comprehensive resource of the HGSC methylome, comprising 404 whole-genome bisulfite sequenced samples from 125 patients belonging to the real-world DECIDER trial. Tumor-specific methylomes were integrated with matched RNA sequencing, whole-genome sequencing and clinical outcomes, which enabled identification of > 8,500 regulatory promoters. Our results show that the regulatory methylome is largely established early in tumor progression and remains stable through chemotherapy. Tumors with poor response to treatment are marked by widespread promoter hypermethylation, particularly cancer cells in ascites, leading to epigenetic silencing of pathways targeted by chemotherapy agents. Our proof-of-principle analysis shows that these epigenetic signatures are detected in plasma-derived DNA, highlighting the value of DNA methylation in liquid biopsy-based disease monitoring. This study provides a foundational resource for dissecting the spatial and temporal dynamics of cancer methylomes and establishes a framework for integrating epigenetic states with clinical trajectories in HGSC.

## Introduction

Ovarian high-grade serous carcinoma (HGSC) is the most lethal gynecologic malignancy, accounting for the majority of ovarian cancer–related deaths (*1*). The high mortality rate is largely due to the late-stage diagnosis, when tumors have already disseminated throughout the peritoneal cavity (*2*), and to the widespread development of resistance to platinum–taxane chemotherapy. Maintenance therapies, such as bevacizumab and poly (ADP-ribose) polymerase (PARP) inhibitors, have improved outcomes in homologous recombination deficient (HRD) tumors (*3, 4*), but most patients ultimately relapse with chemotherapy resistant disease. Thus, there is an urgent need to understand the biological mechanisms driving tumor dissemination and treatment response.

Large-scale sequencing studies have mapped the genomic landscape of HGSC, revealing ubiquitous *TP53* mutations (*5*), extensive copy number alterations (*6*–*8*), and diverse clonal trajectories (*9, 10*). While these studies have assessed the role of genomic aberrations in HGSC, they were not able to fully account for site-specific adaptations or emergence of chemoresistance, suggesting that non-genetic alterations play an important role in HGSC metastasis and chemotherapy resistance.

DNA methylation is an epigenetic reprogramming mechanism that regulates gene expression and cellular identity. It is also crucial in driving tumor adaptation to metastatic niches and chemotherapy resistance through tumor–microenvironment interactions (*11, 12*). As DNA methylation is tissue-specific and stable, it has emerged as a biomarker in clinic routine, including tumor classification (*13, 14*), early detection (*15, 16*), tracing the origin of metastases (*17*) and as an indicator for targeted therapies (*18, 19*). The advent of whole-genome bisulfite sequencing (WGBS) has enabled unbiased, single-base resolution profiling of DNA methylation, providing unprecedented insights into how epigenetic alterations shape cancer development and progression (*20*–*22*). However, in several cancers, such as HGSC, the role of DNA methylation in disease progression and chemotherapy response remains largely unexplored, as existing studies are limited in size (*23*) and confine to a single tumor site or restricted to a list of candidate genes (*24*).

Herein, we characterized the epigenetic landscape of HGSC at single-base resolution of 125 patients with HGSC enrolled to the real-world DECIDER trial (*9*) before and after treatment with WGBS. Integration of transcriptomics, genomics, circulating tumor DNA, and clinical data, along with the decomposition of the DNA methylation to overcome cancer cell fraction variability, enabled comprehensive reconstruction of HGSC regulatory methylomes across space and time. Through this approach, we comprehensively define the DNA methylation landscape of metastatic dissemination and treatment-induced epigenetic changes in HGSC.

## Results

### Multi-site longitudinal DNA methylation landscape of HGSC

The study cohort comprised 125 patients with HGSC enrolled in the prospective, longitudinal, multi-regional DECIDER trial (NCT04846933, **Supplementary Table 1, Methods**). In total, we collected 404 samples from diverse anatomical sites, including primary sites (fallopian tubes, ovaries, uterine adnexa not otherwise specified), intraperitoneal and extraperitoneal metastases, and ascites **(Figure 1a, Supplementary Table 2**). The patients were treated according to the HGSC standard-of-care guidelines (*3*), which consist of surgery and platinum-taxane chemotherapy with angiogenesis and/or PARP inhibitors as maintenance therapy. Samples were collected before chemotherapy (70%), after neoadjuvant chemotherapy (NACT, 19%), and during the treatment of relapsed disease (11%). Methylome profiles were quantified using WGBS with a median coverage of 13× (range: 9×–27×). We also utilized matched RNA sequencing (RNA-seq, 388 samples), whole-genome sequencing (WGS, 375 samples), circulating tumor DNA (ctDNA) methylation sequencing, and clinical outcome variables (**Methods**).

**Figure 1.**
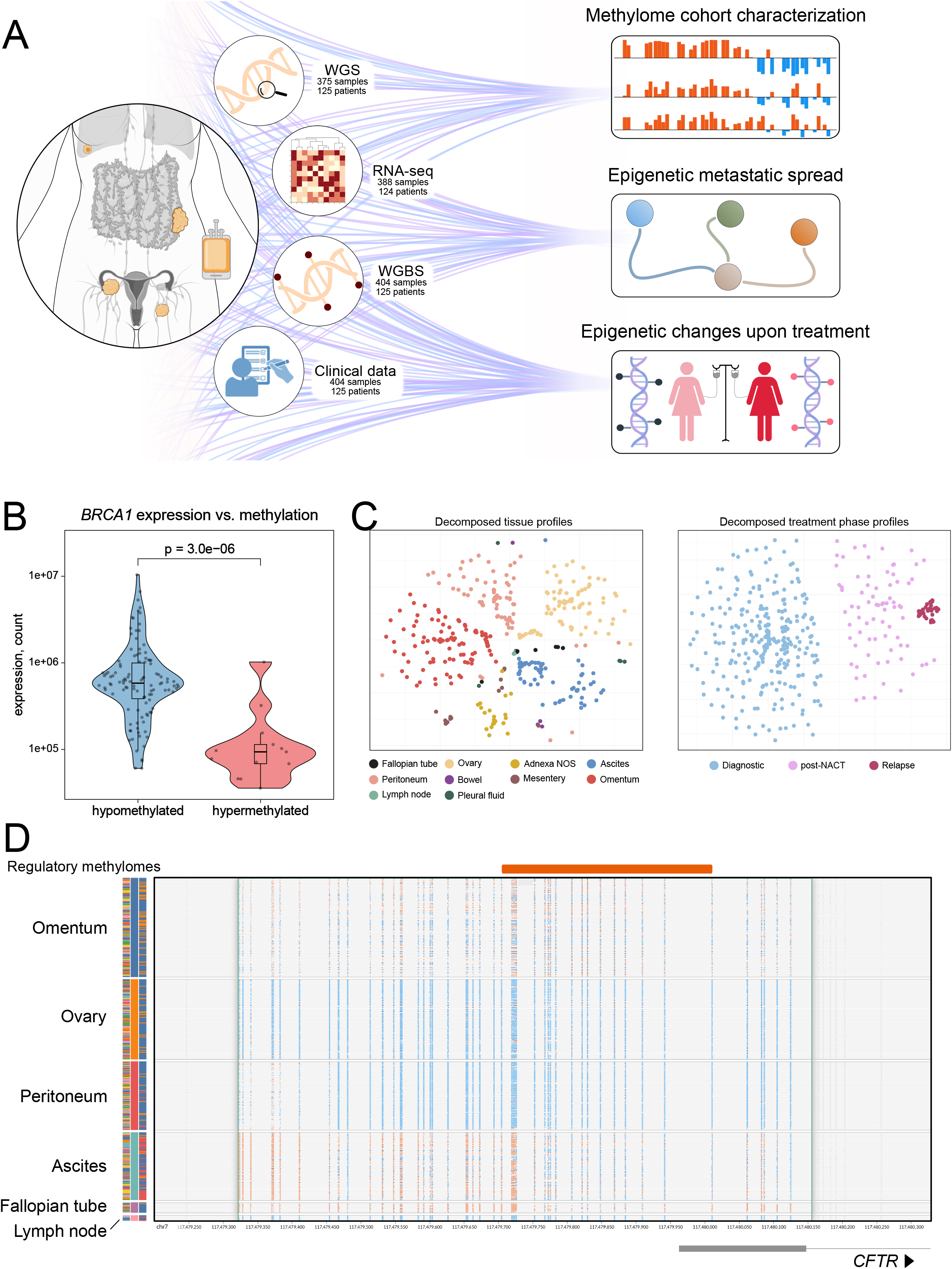
Study workflow and overview of data used in the study: **a**. Study design and schematic summary of data levels used in the three parts of the study. **b**. *BRCA1* promoter hypermethylation was significantly associated with loss of *BRCA1* expression. Data are shown as boxplots and p-value from Mann-Whitney U test is reported. **c**. qSNE plots of ten decomposed tissue (left) and three decomposed treatment phases (right) regulatory methylomes calculated from 404 samples. **d**. A snapshot from GenomeSpy visualization shows DNA methylation of the regulatory region of *CFTR* (orange block), stratified by tumor sites. Each row is one sample. Red and blue bars correspond to hyper- and hypomethylated CpG sites, respectively.

To overcome the bias stemming from varying tumor purity in surgical samples and from patient-specific effects, we used a latent stochastic decomposition model (*25*) for the methylome profiles in all tumor and 18 control samples. To identify regulatory regions, *i*.*e*., CpG loci regulating transcription in cancer cells, we integrated decomposed methylomes and matched decomposed RNA-seq data (**Methods**). This resulted in 8,597 unique promoters, whose methylation levels were significantly inversely correlated with gene expression and are subsequently referred as regulatory methylomes (Spearman correlation, FDR-adjusted *p* < 0.05). For example, hypermethylation of *BRCA1*, which plays a central role in DNA damage repair and is a predictive biomarker for PARP inhibitors (*26*), is strongly associated with the loss of *BRCA1* expression, as shown in **Figure 1b**. All 8,597 regulatory methylomes displayed as qSNE plots (*27*) indicate clear tissue- and treatment phase specificity (**Figure 1c**).

To enable dynamic exploration of this extensive methylome dataset, we have established a GenomeSpy visualization that allows interactive access to per-sample CpG β-values across all 404 samples: https://csbi.ltdk.helsinki.fi/p/marchi_et_al_2025/WGBS.html. The GenomeSpy visualization (*28*) provides quick assessment of methylation patterns across the genome and facilitates exploration of their associations with tumor site and treatment phase. For instance, the *CFTR* decomposed regulatory region displays hypermethylation in ascites and fallopian tube samples, whereas it is hypomethylated in the samples from ovary and peritoneum **(Figure 1d**).

### Regulatory methylome is established early and remains stable through chemotherapy

The distribution of DNA methylation levels within promoter regions provides biologically meaningful insights into the gene regulation (*29, 30*). In normal cellular contexts, unimodal hypomethylation patterns enable consistent transcriptional activation, whereas bimodal distributions, commonly observed in cancers, reflect regulatory heterogeneity and tissue-specific methylation events (*31, 32*). We hypothesized that regulatory promoter regions with varying methylation distributions inform genes that are crucial for cancer progression and treatment response.

To quantify methylation distributions in the 8,597 regulatory methylomes, we used a two-component beta mixture model (**Methods**) and observed that 2.3% (201 genes) exhibited significant bimodal pattern (FDR-adjusted *p* < 0.05). An example of a gene with a bimodal promoter is the *BRCA1* gene (**Figure 2a**). We observed *BRCA1* hypermethylated promoter and loss of gene expression in 13 patients (10.4%), which aligns with the 11% prevalence reported earlier (*33*). As expected, *BRCA1* hypermethylation is significantly associated with HRD (*p* = 4.8e-05, **Figure 2b**) and is mutually exclusive with the genetic loss (*p* = 0.02, **Figure 2c**).

**Figure 2.**
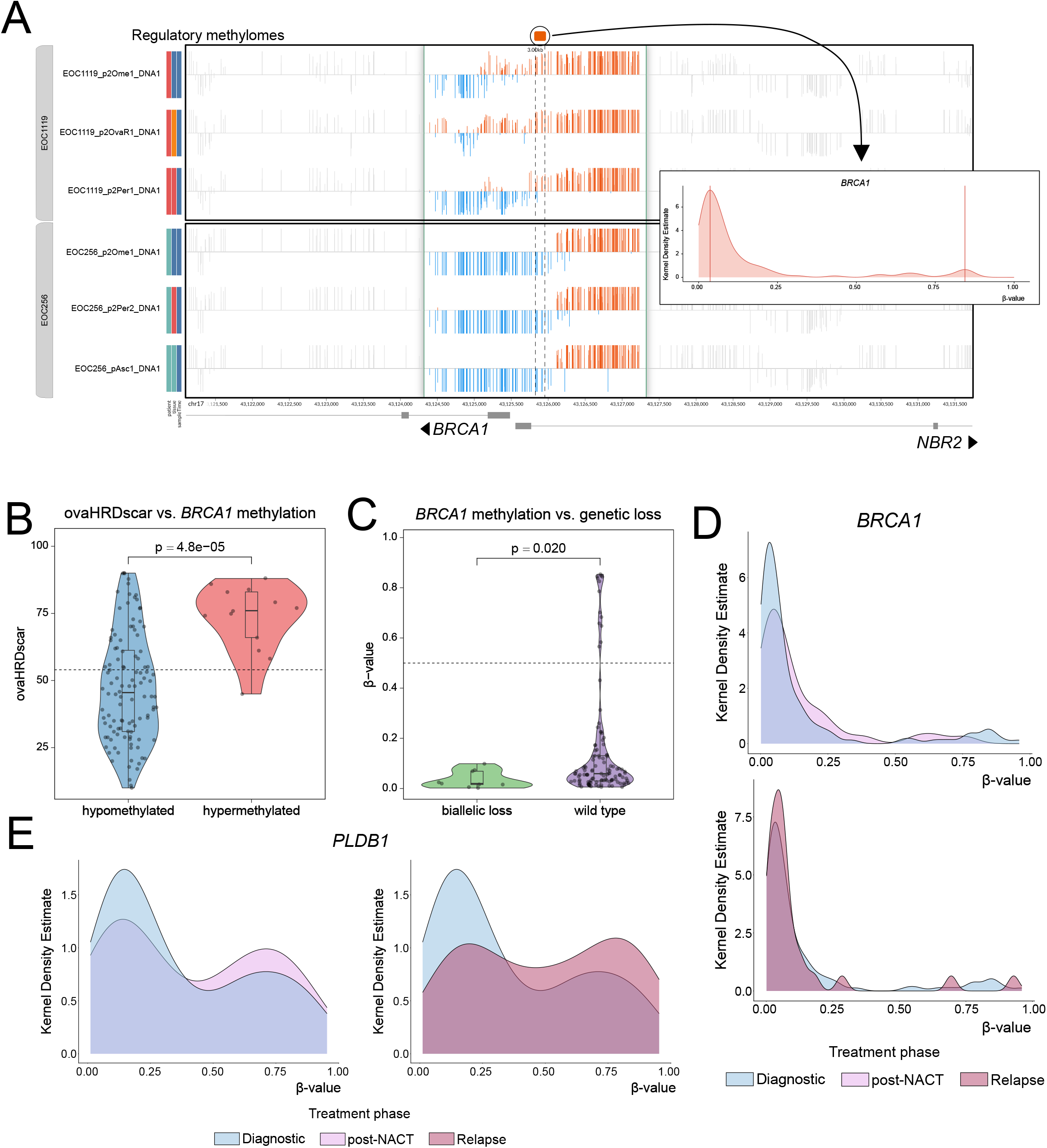
Characterization of the regulatory methylomes: **a**. A snapshot from GenomeSpy visualization shows DNA methylation data of the *BRCA1* promoter from six samples of two patients. Three samples from patient EOC1119 exhibit partial hypermethylation (three top rows), whereas three samples from patient EOC256 are unmethylated. A histogram (right) depicts distribution of beta values across the entire cohort, with a major unmethylated peak and few additional peaks at hypermethylation. **b**. *BRCA1* promoter hypermethylation was significantly associated with HRD-status assessed by ovaHRDScar. Data are shown as boxplots and p-value from Mann-Whitney U test is reported. **c**. *BRCA1* promoter hypermethylation was mutually exclusive with the genetic loss of *BRCA1*. Data are shown as boxplots and p-value from Mann-Whitney U test is reported. **d**. Beta value distribution for the early established bimodal regulatory region of *BRCA1* across entire cohort that remained unchanged after NACT (upper) and at relapse (lower). **e**. Beta value distribution for the late established bimodal regulatory region of *PLDB1* across entire cohort that changed significantly after NACT (left) and at relapse (right).

We then characterized the evolution of the regulatory methylomes’ distributions between the site-of-origin and metastatic samples for the 201 bimodal genes. Importantly, we identified 120 genes whose DNA methylation patterns were established early in the evolution and maintained a stable distribution from primary sites to the intraperitoneal metastatic deposits. Most of the bimodal genes exhibited stable methylomes across treatment phases, remaining unchanged after NACT (67%) and at relapse (63%), as exemplified by *BRCA1* (**Figure 2d, Supplementary Table 3**). Interestingly, genes displaying methylome alterations during metastatic dissemination also underwent changes during chemotherapy treatment (73%), as exemplified by *PLBD1* (**Figure 2e, Supplementary Table 3**). These results support the importance of the regulatory methylome in cancer dissemination and chemotherapy resistance.

### Regulatory methylomes quantify metastatic trajectories in HGSC

To characterize how methylation patterns vary across tumor sites, we clustered site-specific decomposed profiles from 404 samples (**Figure 3a**). Methylomes from sites-of-origin closely resembled those of intraperitoneal solid sites, including omentum, peritoneum, as well as ascites, whereas extraperitoneal metastases, such as pleural effusions and lymph nodes, displayed clearly distinct methylome profiles. To quantify metastatic spread across tumor sites (**Supplementary Table 2**), we performed pairwise differential methylation analyses among 12 tumor sites and estimated inter-site similarity (distance index, see **Methods**) as the proportion of significantly differentially methylated regulatory regions in each pairwise comparison. The similarities between tumor sites ranged from 2.9% (pelvic peritoneum vs. omentum) to 27.3% (pelvic mesentery vs. upper-abdominal peritoneum), with 66 pairs exhibiting distance index < 10% (**Figure 3b)**. Next, using these pairs, we estimated the trajectories from the primary site to the intra- and extraperitoneal metastatic deposits by minimizing distance indices and assessing significance using permutation test. We then quantified the contributions of each trajectory by calculating the fraction of pairs where one of the tumor sites is a) ascites, b) para-aortic lymph node, and c) intraperitoneal metastatic site, of 66 pairs that comprised the model. These analyses resulted in three trajectories of spread: direct dissemination from primary sites to the intraperitoneal metastases (57%), ascites-mediated spread (30%), and lymphatic spread (13%), as shown in **Figure 3c**. Our results align with the established model of intraperitoneal dissemination of HGSC, where major spreading route includes direct dissemination of cancer cells from the primary tumor to neighbor organs within the abdominal cavity (*34*), as well as travelling in the form of spheroids to distant locations (*35*).

**Figure 3.**
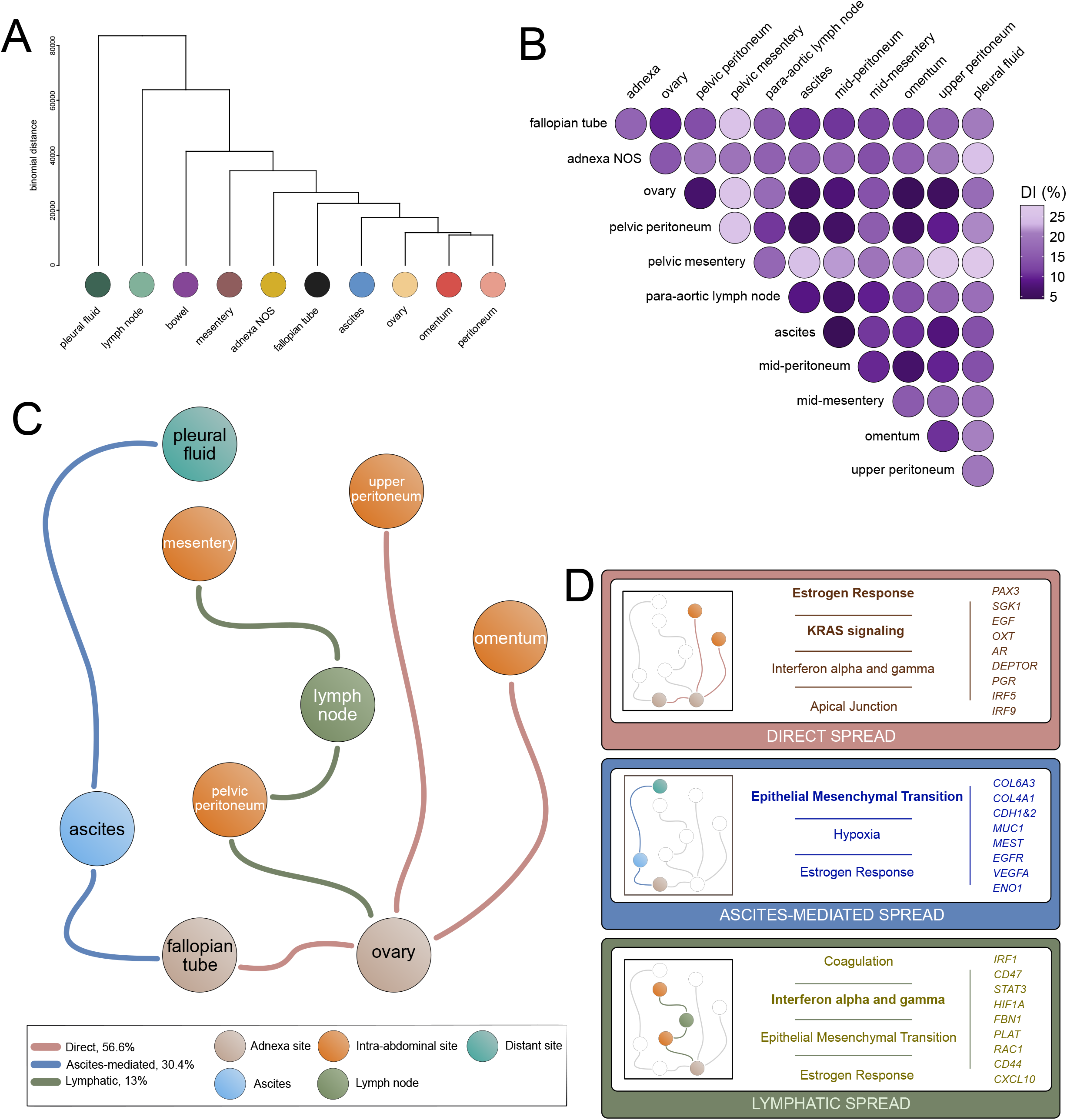
Trajectories of metastatic spread in HGSC: **a**. Hierarchical clustering of 404 decomposed tissue methylomes. The tissues-of-origin tube, ovaries and adnexa NOS co-cluster with ascites, whereas distant metastases form separate branches. **b**. Pair-wise distance matrix of twelve decomposed tumor sites. Smaller (darker) distance index (DI) corresponds to higher similarity between the sites. The smallest DI = 2.9% refers to pelvic peritoneum-omentum, whereas the largest observed DI = 27.3% was observed for comparison pelvic-mesentery – upper-abdominal peritoneum. **c**. Model of spread calculated based on distance indexes revealed three metastatic trajectories stemming from tissues-of-origin. Direct spread (brown links) originated from ovaries to intra-peritoneal metastatic sites (pelvic peritoneum and omentum); ascites-mediated trajectory (blue links) originated from tube and via ascitic circulation reached distant metastases in lungs; lymphatic trajectory (green) stemmed from ovaries via para-aortic lymph nodes reached mesentery. **d**. Characterization of oncogenic pathways underlying metastatic trajectories. Statistically significant pathways (MSigDB Hallmark 2020) enriched in the trajectory at the threshold of FDR-adjusted p < 0.05 are presented per trajectory, with the most significant pathway marked in bold.

To characterize pathways underlying the metastatic spread, we performed pathway analyses using differentially methylated genes for each trajectory (**Methods, Figure 3d, Supplementary Table 4**). The direct peritoneal spread was primarily associated with activation of KRAS and estrogen response, consistent with epigenetic reprogramming that may reflect interactions between tumor cells and the surrounding microenvironment. The ascites-mediated route to pleural cavity showed prominent involvement of epithelial mesenchymal transition (EMT), suggesting that enhanced cell motility contributes to tumor spread via fluids. The lymphatic trajectory was dominated by interferon signaling, supporting an immune-modulated progression along lymphatic channels.

### Chemotherapy induces tissue-specific DNA methylation changes

To evaluate chemotherapy-induced changes in regulatory methylomes, we conducted differential methylation (DM) analyses comparing pre- and post-treatment samples after neoadjuvant or adjuvant treatment (**Methods, Supplementary Table 5**). The impact of NACT on methylomes was modest, with pre- and post-NACT samples differing by only 5% of DM genes and 217 genes out of all 298 genes were hypermethylated, and estrogen signaling emerging as the only significantly enriched pathway. In contrast, relapse samples displayed pronounced alterations (28% DM), affecting key oncogenic and stress-response processes. Notably, promoter hypermethylation resulted in silencing of immune-related genes, while EMT was associated with loss of DNA methylation, suggesting that HGSC recurrence is characterized by epigenetic shifts that suppress the immune response while enhancing tumor migratory potential.

To explore how individual tumor sites respond to therapy, we compared decomposed methylomes from ten sites at diagnosis and after NACT, and from six sites at diagnosis and at relapse. The largest change in the regulatory methylome after NACT occurred in omentum (39.3%, **Figure 4a**), whereas in the relapse samples the largest shift was observed in ascites (54.2%, **Figure 4b**). Thus, while the overall impact of NACT on methylomes is modest, at relapse we observed extensive methylome changes

**Figure 4.**
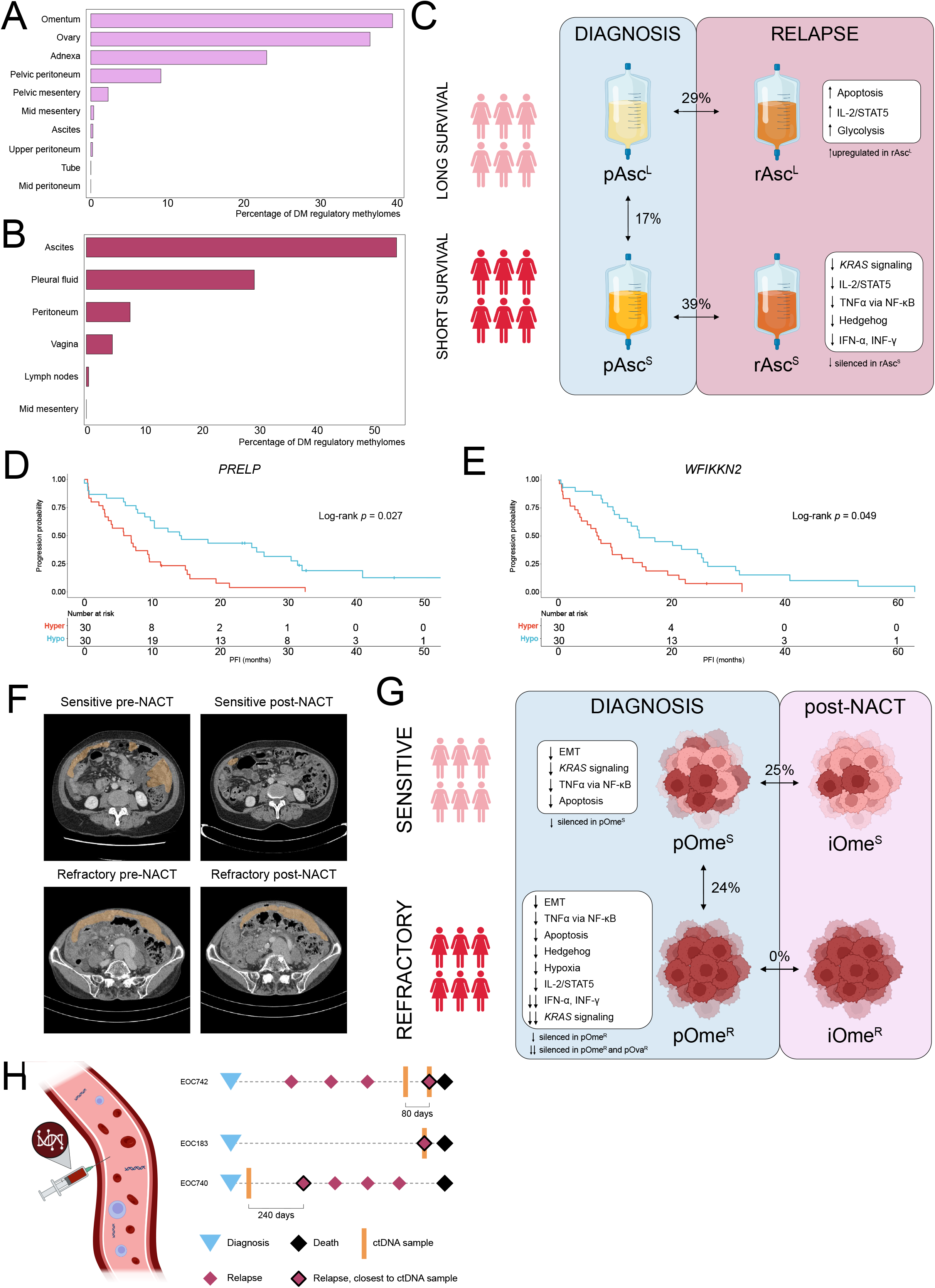
Role of methylome in treatment response: **a**. Tissue-specific changes in decomposed methylomes of ten tumor sites after NACT, expressed as differentially methylated regulatory regions. **b**. Tissue-specific changes in decomposed methylomes of six tumor sites at relapse, expressed as differentially methylated regulatory regions. **c**. Comparison of the ascites methylomes at diagnosis and at relapse in groups of long-(pAsc^L^ vs rAsc^L^) and short-term (pAsc^S^ vs rAsc^S^) survivors. Top statistically significant pathways enriched in the relapse samples of long- and short-term survivors, along with the directions, are presented in the boxes. **d**. Kaplan-Meier platinum-free interval (PFI) curves for 60 patients, stratified into hyper and hypomethylated groups based on beta-value of regulatory region of *PRELP*. **e**. Kaplan-Meier PFI curves for 60 patients, stratified into hyper and hypomethylated groups based on beta-value of regulatory region of *WFIKKN2*. **f**. CT scans of omentum from a good responder (top row) demonstrating a marked reduction in tumor burden (brown) and from a chemo-refractory patient (bottom row) showing minimal change over the treatment course, before and after NACT. **g**. Comparison of the omentum methylomes before and after NACT in groups of sensitive (pOme^S^ vs iOme^S^) and chemo-refractory (pOme^R^ vs iOme^R^) patients. Top statistically significant pathways enriched in the untreated samples of sensitive and chemo-refractory patients, along with the directions, are presented in the boxes. **h**. Analysis of the ctDNA methylation from samples of partial responders. Plasma samples were collected from three patients at timepoints preceding relapse (80, 0, and 240 days prior to progression depicted at timelines).

### Epigenetic reprogramming in ascites shapes therapy response

Recent studies have suggested that long- and short-term survival in HGSC is not dependent on major genetic drivers (*36, 37*). Thus, we hypothesized that epigenetic mechanisms might contribute to differences seen in treatment responses. Given that ascites obtained at relapse exhibited the most extensive methylome changes as compared to diagnostic samples (**Figure 4b**), we characterized the role of ascites in treatment response.

We analyzed ascites samples from 22 short-term (PFI < 6 months) and 10 long-term (PFI > 12 months) survivors by comparing their regulatory methylomes at diagnosis and at relapse (**Figure 4c**). There were substantial changes in both short- and long-term survivors, with 39% and 29% DM regions after treatment, respectively. Hypermethylation of the regulatory regions was abundant at relapse for both short- (64%) and long-term (80%) survivors, indicating global silencing of gene promoters during HGSC progression. Pathway analyses revealed that, particularly in short-term survivors, chemotherapy induced extensive promoter silencing affecting key hormonal and immune signaling networks (**Supplementary Table 6**). These findings suggest that tumor cells in ascites undergo widespread methylome reprogramming that contributes to chemotherapy failure.

Given the observed changes in ascites at relapse, we next investigated its prognostic potential for tracking tumor progression and duration of response. Hence, we compared ascites samples at diagnosis, between short- and long-term survivors and found 17% difference (209/1049 DM genes silenced in short-term survivors). Surprisingly, no pathways reached significance in enrichment analyses, implying that there is no major oncogenic processes underlying intrinsic chemoresistance that are governed by DNA methylation changes. We identified 38 regulatory regions with bimodal methylation patterns that were differentially methylated in short-term survivors (**Supplementary Table 7**). Among these, five showed a significant association between promoter methylation and PFI in samples obtained at diagnosis (log-rank test, *p* < 0.05) of which *PRELP* (FDR-adjusted *p* = 0.027, **Figure 4d**) and *WFIKKN2* (FDR-adjusted *p* = 0.049, **Figure 4e**) were hypermethylated in short-term survivors and remained significant after multi-hypotheses correction. Downregulation of *PRELP* has been found to facilitate cancer cell migration via reduced cell-cell adhesion (*38, 39*) and *WFIKKN2* has been linked to TGF-β– mediated EMT (*40*), supporting the notion that crosstalk between cancer cells and the extracellular matrix contributes to treatment resistance.

### Chemo-refractory tumors show widespread hypermethylation

Similar to the poor- and long-term survival patients, intrinsic chemo-refractory patients, *i*.*e*., patients who do not respond or show minimal response to chemotherapy (**Figure 4f**), have not been found to harbor major genetic drivers (*41, 42*). To investigate whether innate resistance is reflected in the methylomes, we analyzed omental samples from seven chemo-refractory and 27 chemo-sensitive patients collected before and after NACT. The methylomes of chemo-refractory patients remained largely stable (**Figure 4g**), whereas chemo-sensitive patients exhibited substantial alterations, with 25% of regions differing after treatment (834/1110 hypermethylated after NACT). In sensitive patients, NACT induced promoter methylation shifts in pathways regulating cell plasticity and survival (**Supplementary Table 8.A**).

To delineate innate epigenetic differences at diagnosis, we next compared omental methylomes before treatment from chemo-refractory and chemo-sensitive patients. The analysis revealed a 24% difference, driven primarily by widespread hypermethylation in the chemo-refractory group. Strikingly, enriched pathways featured EMT, hypoxia, apoptosis, p53 pathway, as well as several signaling cascades driving tumor progression (**Supplementary Table 8.B**). These results imply that innate chemo-refractory disease is marked by widespread transcriptional silencing of key oncogenic programs, which prevents the use of standard anti-cancer therapies.

To investigate whether the widespread hypermethylation is established already at primary sites, we compared ovarian samples between chemo-sensitive and chemo-refractory patients and found 21% difference (1021/1155 hypermethylated in chemo-refractory samples, see **Supplementary Table 8.C**). Notably, almost one-third of pathways silenced in omentum exhibited hypermethylation already at primary site, indicating that they are established early in the evolution. Thus, our results suggest that in chemo-refractory patients, intra-abdominal dissemination of HGSC prior to treatment is accompanied by extensive epigenetic alterations affecting oncogenic signaling.

As our results from the poor responding patients showed that tumors displayed extensive hypermethylation, we wanted to test whether this epigenetic phenotype is detectable in liquid biopsy samples. Thus, we conducted a proof-of-principle analysis of ctDNA methylation in four samples from three patients with partial response to standard-of-care. The samples were collected at time-points preceding relapse (0, 80, and 240 days prior to relapse) as shown in **Figure 4h** and are available for exploration via GenomeSpy: https://csbi.ltdk.helsinki.fi/p/marchi_et_al_2025/ctDNA.html. We focused on a set of 356 genes hypermethylated in both ovarian and omental samples from chemo-refractory patients, testing whether this epigenetic signature was present in ctDNA during treatment and at relapse. Twenty-three genes showed progressive accumulation of promoter hypermethylation that correlated with decreasing time to relapse (**Supplementary Table 9**), supporting the potential of methylome profiling for monitoring minimal residual disease.

### Discussion

Using a unique whole-genome dataset of 404 samples from 125 patients with HGSC, we characterized the spatial and temporal landscape of the regulatory methylome. Integration of DNA methylation with matched genomic and transcriptomic profiles, together with a latent decomposition, enabled isolation of cancer cell–specific profiles and intra-patient and longitudinal analyses. At the cohort level, the regulatory methylome exhibited extensive intra-patient heterogeneity, and chemotherapy-induced selection, emphasizing the need for multi-site sampling to inform effective therapeutic strategies in HGSC.

Our results based on cancer cell-specific profiles allowed delineation of the relative contribution of the principal routes of metastatic dissemination. Nearly 90% of the spread occurred through direct surface contacts and ascitic flow, in agreement with prior models of HGSC dissemination (*34, 43*). Ascites-mediated trajectory strongly contributed to the dissemination beyond the abdominal cavity, underscoring critical role of ascites in facilitating broader tumor spread, as suggested earlier (*44*–*46*).

Multi-site, longitudinal sampling enabled evaluation of global and site-specific epigenetic responses to neoadjuvant and complete first-line chemotherapy. Regulatory methylomes remained largely stable during neoadjuvant chemotherapy (NACT), whereas relapses were characterized by profound alterations in DNA methylation. Distinct tumor sites exhibited markedly different responses to treatment, ranging from minimal changes in the peritoneum, mesentery, and lymph nodes to extensive remodeling in ascites and omentum. Cancer cell methylomes in ascites showed little change following NACT but diverged markedly at relapse, highlighting substantial epigenetic heterogeneity at recurrence. Genes within EMT and hypoxia pathways were epigenetically silenced, suggesting that these changes provide a selective survival advantage for cancer cells within ascitic fluid under therapeutic pressure. Also, omentum of chemo-sensitive patients showed prominent methylome changes, whereas chemo-refractory patients displayed a constitutively hypermethylated omentum and lacked significant methylation changes. This intrinsic hypermethylation was associated with widespread transcriptional downregulation of key oncogenic and inflammatory pathways, suggesting a repressed regulatory state that may limit responsiveness to most chemotherapies.

To evaluate the degree of inter-patient heterogeneity, we applied a beta mixture model and characterized the methylome distributions in the regulatory regions across the cohort. Earlier studies have shown that gene promoters remain unmethylated in non-malignant cells, whereas in cancer DNA methylation patterns undergo major alterations, resulting in bimodal distribution (*11*). In line with the recent study of non-small cell lung cancer evolution (*47*), we showed that both metastatic spread and poor response to chemotherapy are fueled by marked hypermethylation of regulatory regions. Intra-peritoneal and distant metastatic sites were characterized by silenced methylomes, suggesting that targeting DNA methylation mechanisms may result in a diminished metastatic potential in HGSC. In addition, chemotherapy treatment was associated with an increase of hypermethylation events globally, indicating that cancer cells use DNA methylation to mitigate the impact of platinum-taxane treatment.

We acknowledge limitations of our study. While our cohort is large, some tumor sites, such as lymph nodes and ascites after NACT are represented by 5-10 samples, which may limit the generalizability of the results based on these sites. Moreover, translating the DNA methylation vulnerabilities identified here into effective therapeutic strategies will require validation in independent cohorts.

In conclusion, this study highlights the pivotal role of the methylome in HGSC dissemination and emergence of chemoresistance. Widespread hypermethylation in chemo-refractory patients emphasizes that improvements in HGSC patient care require combinational therapies that should include epigenetic drugs. Leveraging an extensive, longitudinal, multi-site patient dataset, we provide the first comprehensive epigenetic characterization of HGSC metastases and their site-specific responses to treatment.

## Methods

### Sample preparation

#### Solid tissue sample preparation

Tumor tissue samples were obtained from laparoscopy and debulking surgery during normal clinical treatment course and were snap-frozen in LN_2_ within 10 minutes of excision. The Qiagen AllPrep kit (QIAGEN N.V., Germany) was used to extract DNA and RNA simultaneously from the samples.

#### Liquid tissue sample preparation

Whole blood buffy coat (germline) samples were acquired in cooperation with standard clinical care, and the genomic DNA was extracted in Auria biobank using Chemagic DNA Blood Kit Special (PerkinElmer Inc., USA) and Chemagic 360 instrument (PerkinElmer Inc., USA). Ascites and pleural fluid samples were obtained from either laparoscopy and debulking surgery (ascites), paracentesis (ascites), or thoracentesis (pleural fluid) during normal clinical treatment course, and all but two samples were processed within approximately two hours from sample acquisition. Prior to the DNA/RNA extraction, ascites and pleural fluid samples were centrifuged at 1500 rpm for 10 min to precipitate the tumor cells. Cell pellet was resuspended in PBS (phosphate buffered saline), and the cell suspension was pipetted into centrifuge tubes containing Histopaque-1077 density gradient cell separation medium (Sigma Aldrich, Merck KGaA, Germany) to isolate the tumor cells. The tubes were centrifuged at 1500 rpm for 30 min, and the resulting tumor cell isolate layer was pipetted into microcentrifuge tubes and centrifuged at 1500 rpm for 10 min to precipitate the cells. The resulting cell pellet was frozen at −80 °C. The Qiagen AllPrep kit (QIAGEN N.V., Germany) was used to extract DNA and RNA simultaneously from the tumor cell pellets. An additional ethanol precipitation step was performed on one post-NACT ovary sample to purify and concentrate the DNA. Both solid and liquid tumor samples were sent to BGI (BGI Europe A/S, Denmark), where bisulfite conversion was performed on the samples prior to the library preparation and sequencing.

#### ctDNA sample preparation

Blood samples were acquired in cooperation with standard clinical care and further processed by Turku University Hospital Laboratories. In brief, 2 × 10 mL EDTA-coated tubes of whole blood were centrifuged at +4 °C, the plasma layer was separated and then centrifuged again at +4 °C. The twice-centrifuged plasma was divided into 6 parts and frozen at −80 °C within two hours from blood collection. The frozen plasma vials were shipped to BMK (Biomarker Technologies BMK GmbH, Münster, Germany) for cfDNA extraction, which was performed with Quick-cfDNA™ Serum & Plasma Kit (Zymo Research Corporation, Irvine, California) following the standard manual provided by Zymo Research.

### WGBS data processing

WGBS data were processed using Anduril bioinformatics workflow platform (*48*). We trimmed read data using Trimmomatic v0.32 (*49*) and assessed quality with FastQC v0.11.4 (*50*) via Anduril QCFasta v5.0 component. The trimmed reads were aligned to the bisulfite-converted human reference genome (GRCh38.d1.vd1) with Bismark v0.22.3 (*51*), using Bowtie2, allowing one mismatch in the seed (-N 1) and a maximum insert size of 1200 bp for paired-end alignments. The --nucleotide_coverage option was enabled to generate per-base methylation coverage, and unmapped or ambiguous reads were retained and subsequently re-aligned in single-end mode. Deduplication was carried out separately for paired and re-aligned single-end libraries, after which BAM files from multiple sequencing libraries of the same sample were merged using samtools. Alignment and deduplication statistics were compiled with Bismark’s reporting tools. CpG methylation extraction was performed with Bismark’s methylation extractor, using default parameters to avoid counting overlapping mates and trim biased read ends. Cytosine reports were produced in CpG context and further processed with custom R scripts to remove non-standard chromosomes and generate methylation count tables. Coverage and alignment summaries were computed using the Anduril BamStats component, which incorporates BEDTools *genomecov* (*52*). CpG genomic annotations were obtained from the UCSC Genome Browser (hg38).

### WGS data processing

We processed WGS samples as previously described (*9*). Briefly, we trimmed read data using Trimmomatic v0.32 (*49*) and aligned the reads to GRCh38.d1.vd1 using BWA-MEM v0.7.12-r1039. Somatic and germline short variants were called using GATK v4.1.9.0(*53*) Mutect2 and HaplotypeCaller, respectively, in corresponding joint calling approaches. We performed copy-number segmentation using GATK v4.1.4.1 and subsequently estimated copy numbers, purity, and ploidy using a modified version of ASCAT v2.5.2(*54*). We retained samples with purity of at least 10%, resulting in 375 samples from 125 patients.

We estimated homologous recombination deficiency (HRD) with ovaHRDscar (*55*), using copy numbers called as described in (*8*). Samples were considered HRD if their scar score was at least 54. Patients were considered HRD if all samples were HRD and accordingly, patient ovaHRDscar score was the minimum observed for that patient.

### Circulating Tumor DNA Methyl Seq data processing

Circulating tumor DNA (ctDNA) methyl-seq data were available for four samples: EOC740_TPL5583, EOC742_TPL5570, EOC742_TPL8521, and EOC183_TPL8558. Data was processed using the same workflow described in the WGBS data processing section. Briefly, raw sequencing reads were quality controlled, aligned to the hg38 reference genome, and methylation β-values were extracted using the established pipeline.

### WGBS ENCODE data from healthy tissues

As a normal reference, we downloaded 18 WGBS datasets as CpG-level methylation counts from the ENCODE portal (*56*) (https://www.encodeproject.org/) with the following identifiers: ENCFF247ILV, ENCFF318AMC, ENCFF435ETE, ENCFF454NPL, ENCFF477GKI, ENCFF509SPS, ENCFF526FPA, ENCFF553HJV, ENCFF588IUK, ENCFF672QKY, ENCFF689TNG, ENCFF699GKH, ENCFF703XLD, ENCFF716SXG, ENCFF730NQT, ENCFF746GQB, ENCFF953DKC, and ENCFF956ACC. These samples are derived from a variety of healthy tissues, including ovary, adipose, spleen, aorta, muscle, mammary gland, monocytes, B cells, and T cells, and originate from both male and female donors. Male sex chromosome data was discarded. The downloaded CpG methylation counts were used directly in the decomposition analyses without additional preprocessing.

### WGBS *TP53* variant allele fraction estimation

We used the aligned WGBS tumor data to force-call and estimate the variant allele frequencies (VAF) of each patient’s *TP53* variants. To determine callable variants, we used de-converted read data, with T converted C if the mapped reference base on the read’s strand was C or if the read’s base was not mapped to the genome. Similarly, A was converted to G. This data was used to call variants in *TP53* with GATK v4.1.9.0 Mutect2 in joint tumor-only mode, otherwise as described in (*9*). We retained the *TP53* variants that overlapped those as called from WGS data.

Bisulfite conversion of unmethylated C results in reads with C>T or G>A in the reference strand, which biases observed VAF of transition mutations. To account for the conversion, we modeled read counts stratified by strand, *i*.*e*., whether conversion was C>T or G>A in a given read. We obtained strand-specific read counts using SAMtools v1.14 (*57*) mpileup and annotated each read with the strand. Reads were then bucketed by strand and five read types, i.e., base (C, T, G, A) or indel (x; position is an indel or is followed by an indel, which always refers to a variant rather than a bisulfite conversion), resulting in a 10-element vector ***x***. The read counts were modeled with multinomial distribution as

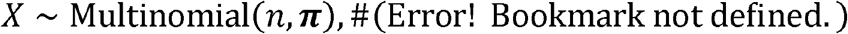

with

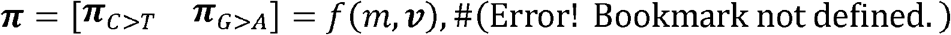

where *n* is the total read count, ***π*** the probabilities of 10 different read types and strands as a function of the local bisulfite conversion rate *m*, and ***ν***, the vector of latent VAFs of bases (C, T, G, A) and indels (x). and are subject to the constraints

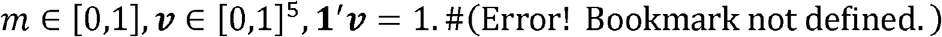

The probabilities for the C>T and G>A strands were defined as

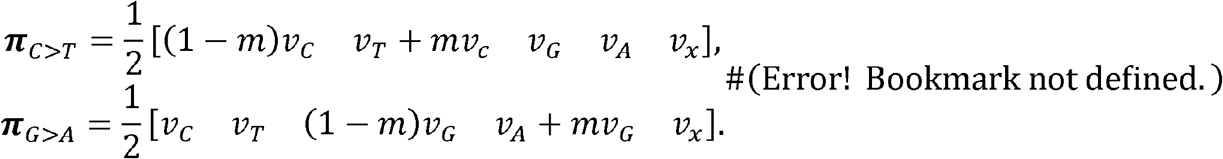

We estimated the parameters and by maximizing the likelihood using Newton’s method with log-barriers for the constraints. The conversion-adjusted *TP53* VAF estimate was defined as the sum of the estimated VAFs of non-reference alleles. For WGBS samples where the *TP53* variant was uncallable, VAF from the corresponding WGS sample was taken. Virtual “purified” bisulfite conversion free reads can be estimated following (*25*), as in Error! Bookmark, and were used to guide the decomposition purity as described below.

### WBGS methylation decomposition

The methylation data are analyzed using a model following (*25*). Specifically, it is assumed that the read counts can be modeled as follows:

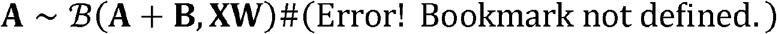

Where 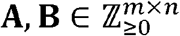 are the methylated and unmethylated read counts, respectively, at the *m* CpG methylation loci in the samples; **X** ∈ [0,1]^*m*×*k*^ represent latent probabilistic methylation profiles at the *m* loci for the *k* profiles, the value representing probability of a methylated read; **W** ∈ [0,1]^*k*×*n*^ represent the profile contributions (the mixing matrix) for the *k* profiles in the *n* samples; and ℬ (*n, p*) is the binomial distribution with draws of probability *p* each.

In our setting, we stack the relevant normal profiles, a patient specific profile, a phase specific profile (diagnosis, after NACT, or relapse), and a tissue specific profile into each sample. These are encoded by an adjacency matrix **M** ∈ {0,1}^*k*×*n*^, which acts as a mask for **W**. Normal samples from ENCODE are augmented to the patient samples to guide the normal profiles (**Supplementary Figure 1**).

Here, are not known, but learned by a decomposition, given the observed read data **A, B**. Due to the sparsity of **W** implied by **M**, it is possible to invert even large systems, as long as the design is nonsingular, that is, no sample features multiple unique profiles to be resolved.

For maximum-likelihood (ML) estimation, the model of (Error! Bookmark not defined.) implies a log-likelihood objective of:

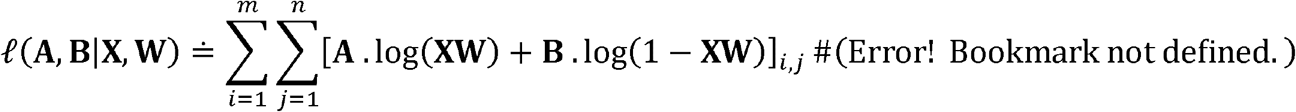

The log-sums can be separated using the variational approximation:

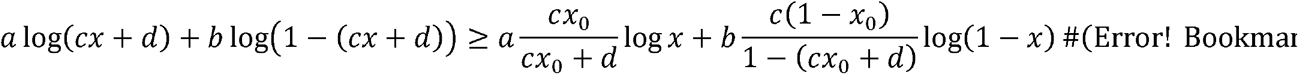

about *x* = *x*_0_(*58*), which is dual to the following expectation–maximization (EM) iteration, here for **X**:

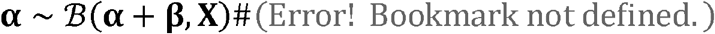

with:

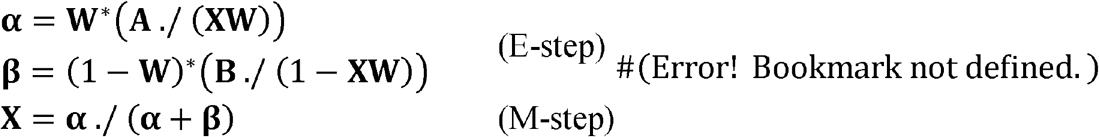

which is just a regular binomial ML stage. A symmetric procedure for **W** exists, however, to constrain **W** to unit mixing weights **1**^**∗**^**W** = 1, the following Lagrangian is constructed:

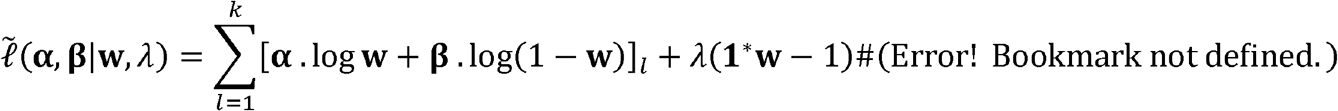

which is optimal at the quadratic root:

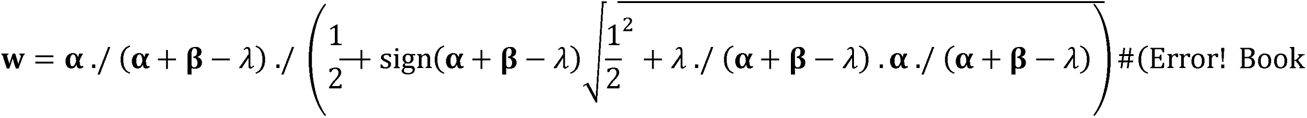

which can be bisected for the multiplier *λ*.

Finally, we can outline the full ML procedure as follows:

1. Choose the initial parameters. We choose **X** = **1**/2, that is 50%/50% un-/methylated profiles, and **W** = **M**./(**1**^**∗**^**M**), that is uniform weights, but the choice is not important if enough data is available and the objective is convex
2. Update **X** using (Error! Bookmark t defined.)
3. Update **W** by computing the moments ***α, β*** from (Error! Bookmark not defined.) with **X, W**^**∗**^ exchanged, and solve 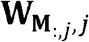 on the nonzero components **M**_:,*j*_ of each sample *j* using (Error! Bookmark not defined.) by choosing *λ*_*j*_ such that **1**^**∗**^**W** = 1
4. Repeat steps 2. and 3. to convergence in (Error! Bookmark not defined.)

It is also possible to solve only for **X** (**W**) if **W** (**X**) is known by omitting step 3. (step 2.), and to construct various mixed mode problems (*i*.*e*., parts of **X, W**are known, parts unknown). In fact, we augment **A, B** with the read counts of selected *TP53* mutations (see the previous section) and **X** with 0s in the normal and 1s in the tumor profiles to guide the purity estimation, as these are known to be clonal.

As described in (*25*), while the profiles **X** can be directly used to compare the methylation levels, the model can be used to decompose the observed read counts **A**, **B** into expected component specific read contributions for post-hoc analysis of “purified” data:

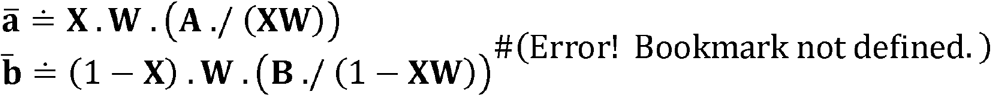

Where 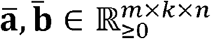 reflect the predicted methylated and unmethylated counts, respectively, at the *m* loci of *k* profiles of *n* samples; or the binomial mode (*59*) can be used for the corresponding maximum posterior estimates (*25*).

Our implementation is available at https://github.com/anthakki/bino, where the bino.fit.*() functions implement the estimators and bino.cond.*() the predictors.

### Annotation of the CpG loci

To characterize CpG loci, we performed gene annotation using GENCODE v40 (hg38). The hierarchical annotation ensured that each CpG site was assigned to a single most relevant genomic element. Gene body regions were given the lowest priority to prevent coding regions from being misclassified as promoters. Upstream, 5′ UTR, and 3′ UTR regions were assigned intermediate priority, reflecting their well-defined structural and regulatory roles. The first exon was assigned the highest priority, as it frequently encompasses promoter-proximal CpGs and exhibits regulatory methylation patterns (*60*). The assignments were strand-specific, with the positions of the first exon and 5′ UTR determined relative to transcript orientation. In the hierarchical scheme described above, higher-priority elements (e.g., first exon > UTRs > gene body) replaced lower-priority ones when overlapping a CpG site. Overlaps between elements of the same category, thus at the same priority level, were marked as *ambiguous* and excluded from further analysis. Each CpG was annotated with gene symbol, gene ID, and strand. Contiguous loci with identical annotations were grouped into unique regional identifiers, yielding 489,739 regions with gene annotation. Annotation was implemented in R v3.6.3 (https://www.r-project.org/).

### RNA-seq processing and integration

Preprocessing and decomposition of 388 RNA-seq samples from 124 patients into cancer, stroma, and immune components were carried out as described in (*58*). Raw counts were normalized for sample-specific sequencing depth, and component contributions were estimated with PRISM(*58*) to obtain component-adjusted expression values. Gene identifiers were harmonized by removing transcript suffixes to maximize overlap with DNA methylation annotations.

The annotated CpG regions (see previous chapter) that mapped to pre-5′ UTR or 5′ UTR elements were considered as putative promoters. Spearman correlation coefficients were computed across matched samples between RNA expression values for the annotated gene and the methylation β-values for each of these putative promoter regions. A promoter was considered regulatory if it showed negative correlation with expression and passed multiple testing correction (BH-adjusted *p* < 0.05) (**Supplementary Figure 2**). This procedure identified 12,760 regulatory methylome–gene associations from 8,597 unique genes.

### Differential methylation testing

In terms of the model of (Error! Bookmark not defined.), testing for differential methylation (DM) at the locus *i* between the component profiles *l*_1_ and *l*_2_ corresponds to testing the null hypothesis 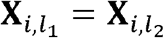. More generally, we can set up a projection **P** ∈ {0.1}*k*′ × *k* from the *k* profiles to a reduced set of *k*′ profiles, which allows more complex profile equivalence, and a relevant set *m*′ loci can be considered for a differentially methylated region instead of a single locus. This is analogous to variance analysis (ANOVA) on the binomial regression of the model of (Error! Bookmark not defined.).

Following (*25*), p-values are computed using the following likelihood ratio test procedure.

1. Project the current model **X**^(1)^ to the appropriate subspace through **X**^(1)^ = **X**^(1)^ **P**^−1^ which essentially averages the profiles in **X** and sums their weights in **W** as **PW**
2. Use the ML estimation procedure to optimize **X**^(0)^
3. Compute the log-likelihoods using with versus (Error! Bookmark not defined.) with **X**^(1)^, **W** versus **X**^(0)^ **PW** and use Wilks’ theorem (*61*) to draw p-values. This assumes −2(*ℓ*^(0)^ − *ℓ1 ~ χ2ν* where *ℓ0, ℓ1* are the null and alternative log-likelihoods, respectively; *v* = *m*′(*k* − *k′*) is the total projection nullity; and *χ*^2^(*ν)* is the chi-squared distribution with *ν* degrees of freedom

Further, one-tailed tests for specifically hypo- or hypermethylation can be realized by summing the log-likelihoods over the sites with the appropriate sign of difference.

Our implementation is available at https://github.com/anthakki/bino where bino.test.proj() computes the projection and the optimized likelihood ratios and bino.wilks() evaluates the p-value.

### Visualizing sample similarity using QSNE

To visualize sample similarity, we use the stochastic neighbor embedding framework popularized by t-SNE (*62*). A regular t-SNE uses a normal input distribution that is to be preserved by the embedding, which is suitable for data that can be easily standardized but is poorly suited to the discrete and heteroscedastic sequencing data. However, this can be mitigated by substituting for the appropriate input distribution.

A binomial density implies the following information gain ratio:

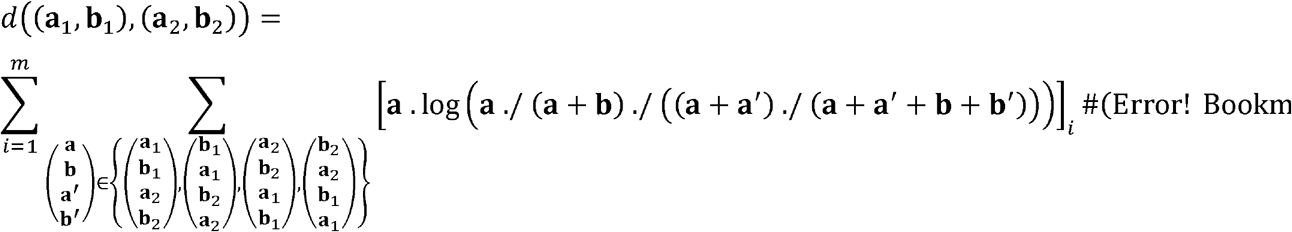

which is the analog of the squared Euclidean distance in preserving binomial information for the purposes of discriminating between the samples **a**_1_, **b**_1_ and 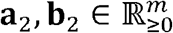.

To compute the embedding, (Error! Bookmark not defined.) is used to compute a pairwise distance matrix between all the samples, which is plugged into a t-SNE optimizer. Either the original data **A, B** or the component predictions, *i*.*e*., 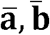 from (Error! Bookmark not defined.), can be used in the process, where the latter enables visualizing patient, treatment phase, and tissue similarities in the absence of normal and the other components.

Our implementation is available at https://github.com/anthakki/bino where bino.dist() implements the distance computation and QSNE (*27*) is used to optimize the embedding.

### Bimodal modeling of regulatory methylomes

To identify genes with bimodal regulatory methylomes’ patterns across patients, we implemented a two-component beta mixture model. For each gene, mean regulatory methylome β-values (methylation at the regulatory promoter) were calculated across samples belonging to the same patient, resulting in a patient-level methylation matrix. We compared the fit of a single-component beta distribution with that of a two-component beta mixture estimated using an expectation–maximization (EM) algorithm. Parameters for each component (α, β) were updated iteratively to maximize the log-likelihood, with mixture weights estimated from posterior probabilities. For each gene, a likelihood ratio statistic was computed, and statistical significance was assessed using a parametric bootstrap procedure under the null hypothesis of a single-component model. Genes were classified as bimodal if the two-component model provided a significantly better fit (bootstrap *p* < 0.05) and the component means were separated by more than 0.5. This framework allowed us to systematically detect genes exhibiting bimodal methylation states across the cohort.

### Hierarchical clustering

To investigate the relationships between tissues, we performed hierarchical clustering using the binomial distance metric defined in Equation (8). The pairwise distance matrix was computed from the tissue-only methylation component. Clustering was performed with the complete linkage method using the R *hclust* function.

### Model of metastatic spread

Distance indexes (DIs), defined as the proportion of significantly differentially methylated regulatory methylomes between two tissue sites, were used to construct a model of metastatic spread across anatomical locations. For each tissue, the full set of DIs against all other sites was obtained, and candidate links were defined as those within a fixed tolerance of ≈1.16% in DI relative to the smallest value, ensuring that only closely related comparisons were retained for further testing. This threshold corresponds to the proportion of 100 genes (100/8,597 ≈ 1.16%) out of the 8,597 regulatory methylomes’ corresponding genes, representing a conservative absolute count large enough to be robust to small sampling fluctuations, yet small enough to exclude clearly distant comparisons. Statistical significance of candidate links was assessed with one-sided permutation tests in which each observed DI was compared against a null distribution generated from 10,000 random permutations of the remaining DI values. P-values were adjusted for multiple testing using BH method to control the false discovery rate. Links were considered significant if they met both criteria: (i) an DI within the 1.16% tolerance of the minimum for the corresponding tissue, and (ii) an adjusted p-value below 0.01.

### Radiological imaging

Computed tomography (CT) imaging was performed as part of standard treatment protocols. Patients underwent contrast-enhanced CT scans, which were reviewed by a single radiologist (JA) with over five years of experience in oncologic imaging.

## Acknowledgements

We are grateful to Dr. Déborah Boyenval for consultation on mathematical modeling. The authors wish to acknowledge the CSC - IT Center for Science (Finland) for computational resources. We thank the patients for their participation, the Turku University Hospital staff for support in recruitment and sampling, and Peppi Alho for technical assistance.

## Authors contributions

Conceptualization: A.L., G.M., K.L., A.V., A.H., J.H., and S.H.; Methodology and Software: G.M., K.L., Y.L., Su.H., and A.H.; Formal Analysis: G.M., A.L., Y.L., V.M.I., D.A., E.P., and J.O.; Investigation: G.M., C.F., T.A.M., F.D., V.M.I., and A.L.; Resources: G.M., A.L., I.M., Y.L., K.L., E.V., Su.H., J.A., D.A., A.H., J.O., J.H., A.V., and S.H.; Data curation: Y.L., V.M.I., T.A.M., J.O., J.H., and A.V.; Visualization: G.M., K.L., Su.H., V.M.I., Y.L., J.A., J.O., A.H., and A.L.; Supervision: A.H., A.L., J.H., A.V., and S.H.; Project Administration: A.L. and S.H.; Funding Acquisition: A.V., A.H., J.H., A.L., and S.H.; Writing Original Draft: G.M.; Writing Review and Editing: all authors.

## Data and materials availability

Whole genome bisulfite sequencing data has been deposited at the European Genome-phenome Archive (EGA), which is hosted by the EBI and the CRG, under accession number EGAS50000000727. Raw WGS and RNA sequencing data are available in the EGA under study accession number EGAS00001006775 and EGAS00001004714, respectively. The implementation of the decomposition, differential methylation analyses, and distance calculation for the qSNE visualization, supplemented by examples, are available at https://github.com/anthakki/bino.

## Supplementary Figures

**Supplementary Figure 1.**
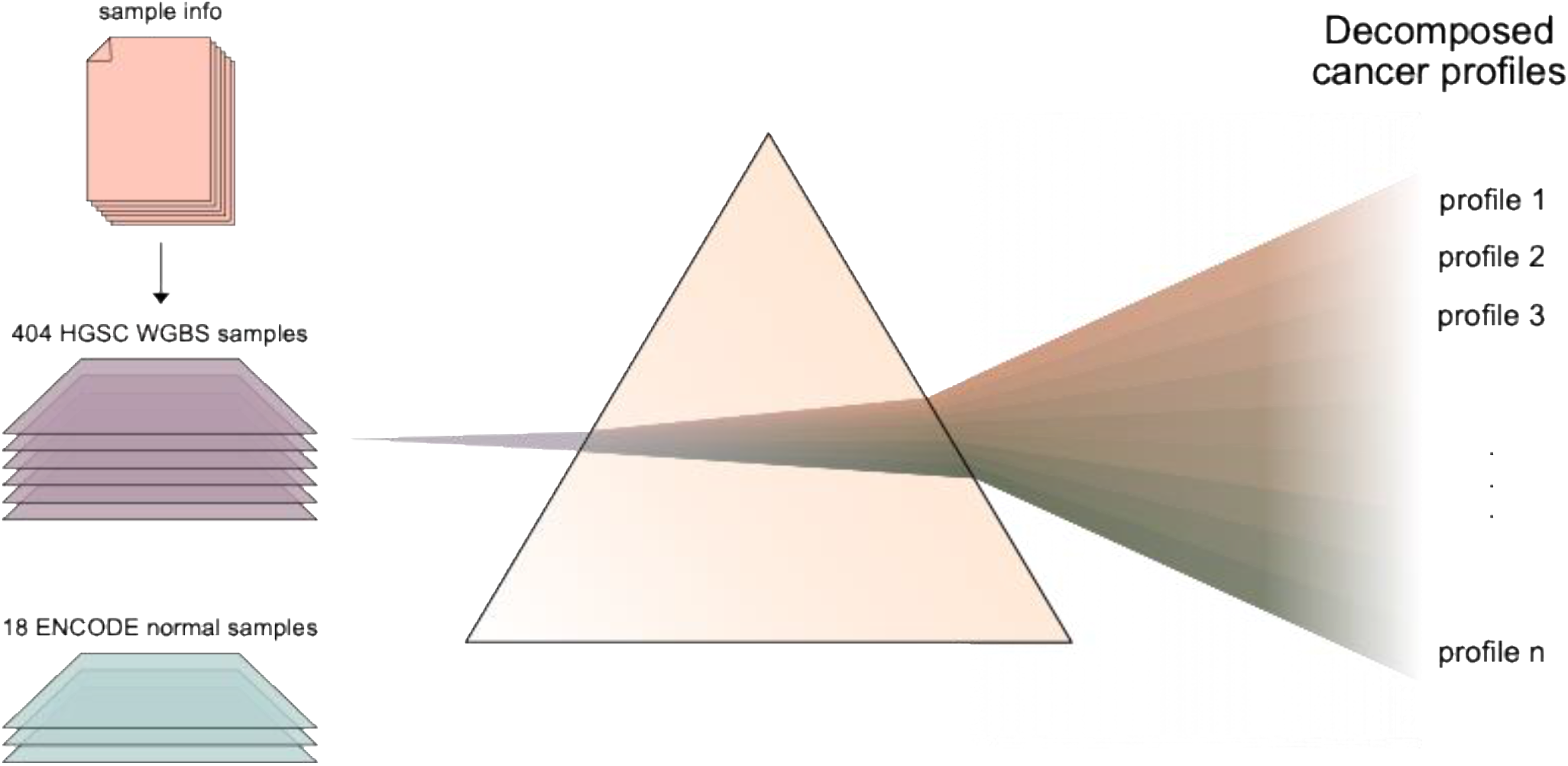
Decomposition of the WGBS data. All 404 cancer samples were used in the decomposition, supplemented by 18 samples from healthy controls. The resulting decomposed cancer profiles of ten tumor sites and three treatment phases were used in the downstream analyses.

**Supplementary Figure 2.**
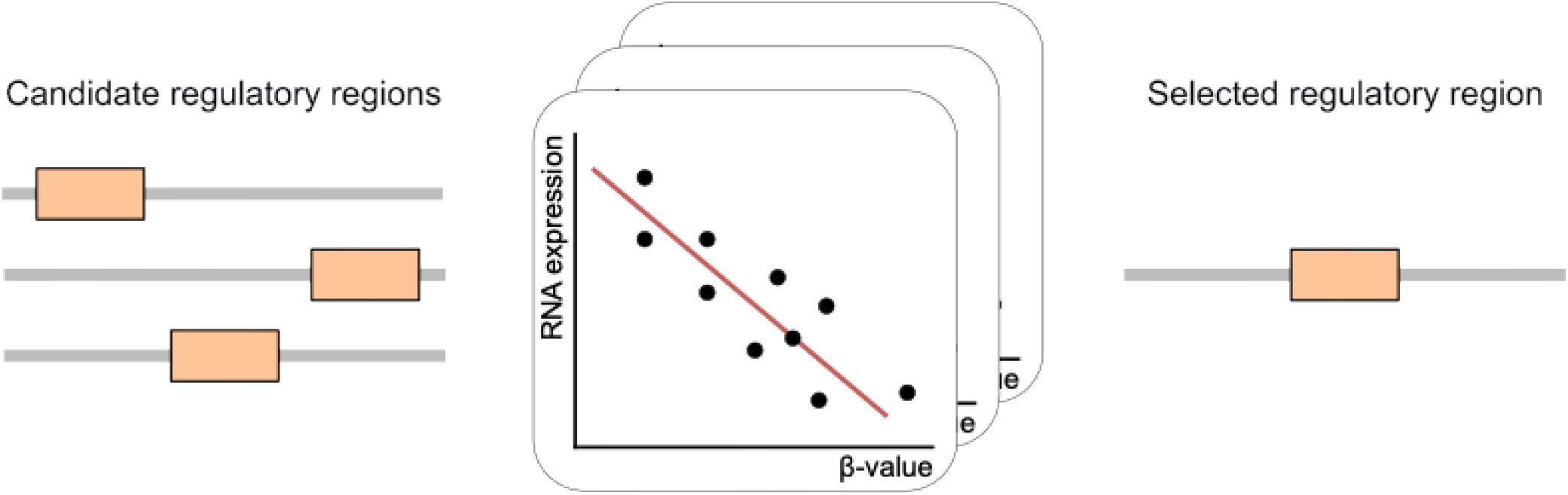
Integration of DNA methylation and matched RNA-seq data. For each gene several putative promoters were identified, which was followed by correlation analyses and resulted in 8,597 unique regulatory regions. Each regulatory region shows significant negative correlation of the beta value with the gene expression.

## References

1. C. J. Cabasag, P. J. Fagan, J. Ferlay, J. Vignat, M. Laversanne, L. Liu, M. A. van der Aa, F. Bray, I. Soerjomataram, Ovarian cancer today and tomorrow: A global assessment by world region and Human Development Index using GLOBOCAN 2020. Int. J. Cancer 151, 1535–1541 (2022).

2. S. I. Labidi-Galy, E. Papp, D. Hallberg, N. Niknafs, V. Adleff, M. Noe, R. Bhattacharya, M. Novak, S. Jones, J. Phallen, C. A. Hruban, M. S. Hirsch, D. I. Lin, L. Schwartz, C. L. Maire, J.-C. Tille, M. Bowden, A. Ayhan, L. D. Wood, R. B. Scharpf, R. Kurman, T.-L. Wang, I.-M. Shih, R. Karchin, R. Drapkin, V. E. Velculescu, High grade serous ovarian carcinomas originate in the fallopian tube. Nat. Commun. 8, 1093 (2017).

3. A. González-Martín, P. Harter, A. Leary, D. Lorusso, R. E. Miller, B. Pothuri, I. Ray-Coquard, D. S. P. Tan, E. Bellet, A. Oaknin, J. A. Ledermann, ESMO Guidelines Committee. Electronic address, Newly diagnosed and relapsed epithelial ovarian cancer: ESMO Clinical Practice Guideline for diagnosis, treatment and follow-up. Ann. Oncol. 34, 833–848 (2023).

4. I. Ray-Coquard, A. Leary, S. Pignata, C. Cropet, A. González-Martín, C. Marth, S. Nagao, I. Vergote, N. Colombo, J. Mäenpää, F. Selle, J. Sehouli, D. Lorusso, E. M. Guerra Alia, G. Bogner, H. Yoshida, C. Lefeuvre-Plesse, P. Buderath, A. M. Mosconi, A. Lortholary, A. Burges, J. Medioni, A. El-Balat, M. Rodrigues, T.-W. Park-Simon, C. Dubot, D. Denschlag, B. You, E. Pujade-Lauraine, P. Harter, PAOLA-1/ENGOT-ov25 investigators, Olaparib plus bevacizumab first-line maintenance in ovarian cancer: final overall survival results from the PAOLA-1/ENGOT-ov25 trial. Ann. Oncol. 34, 681–692 (2023).

5. Cancer Genome Atlas Research Network, Integrated genomic analyses of ovarian carcinoma. Nature 474, 609–15 (2011).

6. S. Jamalzadeh, J. Dai, K. Lavikka, Y. Li, J. Jiang, K. Huhtinen, A. Virtanen, J. Oikkonen, S. Hietanen, J. Hynninen, A. Vähärautio, A. Häkkinen, S. Hautaniemi, Genome-wide quantification of copy-number aberration impact on gene expression in ovarian high-grade serous carcinoma. BMC Cancer 24, 173 (2024).

7. F. C. Martins, D.-L. Couturier, I. de Santiago, C. M. Sauer, M. Vias, M. Angelova, D. Sanders, A. Piskorz, J. Hall, K. Hosking, A. Amirthanayagam, S. Cosulich, L. Carnevalli, B. Davies, T. B. K. Watkins, I. G. Funingana, H. Bolton, K. Haldar, J. Latimer, P. Baldwin, R. Crawford, M. Eldridge, B. Basu, M. Jimenez-Linan, A. W. Mcpherson, N. McGranahan, K. Litchfield, S. P. Shah, I. McNeish, C. Caldas, G. Evan, C. Swanton, J. D. Brenton, Clonal somatic copy number altered driver events inform drug sensitivity in high-grade serous ovarian cancer. Nat. Commun. 13, 6360 (2022).

8. G. Micoli, K. Lavikka, Y. Li, A. Pirttikoski, D. Afenteva, W. Senkowski, G. Marchi, A. Vähärautio, T. A. Muranen, T. Joutsiniemi, S. Hietanen, A. Virtanen, K. Wennerberg, J. Hynninen, J. Oikkonen, S. Hautaniemi, Decoding the Genomic and Functional Landscape of Emerging Subtypes in Ovarian Cancer. Cancer Discov. 15, 2262–2277 (2025).

9. A. Lahtinen, K. Lavikka, A. Virtanen, Y. Li, S. Jamalzadeh, A. Skorda, A. R. Lauridsen, K. Zhang, G. Marchi, V. M. Isoviita, V. Ariotta, O. Lehtonen, T. A. Muranen, K. Huhtinen, O. Carpén, S. Hietanen, W. Senkowski, T. Kallunki, A. Häkkinen, J. Hynninen, J. Oikkonen, S. Hautaniemi, Evolutionary states and trajectories characterized by distinct pathways stratify patients with ovarian high grade serous carcinoma. Cancer Cell 41, 1103–1117.e12 (2023).

10. M. J. Williams, I. Vázquez-García, G. Tam, M. Wu, N. Varice, E. Havasov, H. Shi, D. H. Al-Rawi, G. Satas, H. J. Lees, J. J. K. Lee, M. A. Myers, M. Zatzman, N. Rusk, E. Ali, R. H. Shah, M. F. Berger, N. Mohibullah, Y. Lakhman, D. S. Chi, N. R. Abu-Rustum, C. Aghajanian, A. McPherson, D. Zamarin, B. Loomis, B. Weigelt, C. F. Friedman, S. P. Shah, Tracking clonal evolution during treatment in ovarian cancer using cell-free DNA. Nature 2025, 1–9 (2025).

11. M. Esteller, M. A. Dawson, C. Kadoch, F. V. Rassool, P. A. Jones, S. B. Baylin, The Epigenetic Hallmarks of Cancer. Cancer Discov. 14, 1783–1809 (2024).

12. D. Hanahan, Hallmarks of Cancer: New Dimensions. Cancer Discov. 12, 31–46 (2022).

13. T. Zhu, J. Liu, S. Beck, S. Pan, D. Capper, M. Lechner, C. Thirlwell, C. E. Breeze, A. E. Teschendorff, A pan-tissue DNA methylation atlas enables in silico decomposition of human tissue methylomes at cell-type resolution. Nature Methods 2022 19:3 19, 296–306 (2022).

14. M. C. Liu, G. R. Oxnard, E. A. Klein, C. Swanton, M. V. Seiden, M. C. Liu, G. R. Oxnard, E. A. Klein, D. Smith, D. Richards, T. J. Yeatman, A. L. Cohn, R. Lapham, J. Clement, A. S. Parker, M. K. Tummala, K. McIntyre, M. A. Sekeres, A. H. Bryce, R. Siegel, X. Wang, D. P. Cosgrove, N. R. Abu-Rustum, J. Trent, D. D. Thiel, C. Becerra, M. Agrawal, L. E. Garbo, J. K. Giguere, R. M. Michels, R. P. Harris, S. L. Richey, T. A. McCarthy, D. M. Waterhouse, F. J. Couch, S. T. Wilks, A. K. Krie, R. Balaraman, A. Restrepo, M. W. Meshad, K. Rieger-Christ, T. Sullivan, C. M. Lee, D. R. Greenwald, W. Oh, C. K. Tsao, N. Fleshner, H. F. Kennecke, M. F. Khalil, D. R. Spigel, A. P. Manhas, B. K. Ulrich, P. A. Kovoor, C. Stokoe, J. G. Courtright, H. A. Yimer, T. G. Larson, M. V. Seiden, S. R. Cummings, F. Absalan, G. Alexander, B. Allen, H. Amini, A. M. Aravanis, S. Bagaria, L. Bazargan, J. F. Beausang, J. Berman, C. Betts, A. Blocker, J. Bredno, R. Calef, G. Cann, J. Carter, C. Chang, H. Chawla, X. Chen, T. C. Chien, D. Civello, K. Davydov, V. Demas, M. Desai, Z. Dong, S. Fayzullina, A. P. Fields, D. Filippova, P. Freese, E. T. Fung, S. Gnerre, S. Gross, M. Halks-Miller, M. P. Hall, A. R. Hartman, C. Hou, E. Hubbell, N. Hunkapiller, K. Jagadeesh, A. Jamshidi, R. Jiang, B. Jung, T. H. Kim, R. D. Klausner, K. Kurtzman, M. Lee, W. Lin, J. Lipson, H. Liu, Q. Liu, M. Lopatin, T. Maddala, M. C. Maher, C. Melton, A. Mich, S. Nautiyal, J. Newman, J. Newman, V. Nicula, C. Nicolaou, O. Nikolic, W. Pan, S. Patel, S. A. Prins, R. Rava, N. Ronaghi, O. Sakarya, R. V. Satya, J. Schellenberger, E. Scott, A. J. Sehnert, R. Shaknovich, A. Shanmugam, K. C. Shashidhar, L. Shen, A. Shenoy, S. Shojaee, P. Singh, K. K. Steffen, S. Tang, J. M. Toung, A. Valouev, O. Venn, R. T. Williams, T. Wu, H. H. Xu, C. Yakym, X. Yang, J. Yecies, A. S. Yip, J. Youngren, J. Yue, J. Zhang, L. Zhang, L. (Quan) Zhang, N. Zhang, C. Curtis, D. A. Berry, Sensitive and specific multi-cancer detection and localization using methylation signatures in cell-free DNA. Annals of Oncology 31, 745–759 (2020).

15. Y. Dong, H. Zhao, H. Li, X. Li, S. Yang, DNA methylation as an early diagnostic marker of cancer (Review). Biomed. Rep. 2, 326–330 (2014).

16. J. Liu, L. Dai, Q. Wang, C. Li, Z. Liu, T. Gong, H. Xu, Z. Jia, W. Sun, X. Wang, M. Lu, T. Shang, N. Zhao, J. Cai, Z. Li, H. Chen, J. Su, Z. Liu, Multimodal analysis of cfDNA methylomes for early detecting esophageal squamous cell carcinoma and precancerous lesions. Nature Communications 2024 15:1 15, 1–16 (2024).

17. B. Zhang, Y. Zhou, N. Lin, R. F. Lowdon, C. Hong, R. P. Nagarajan, J. B. Cheng, D. Li, M. Stevens, H. J. Lee, X. Xing, J. Zhou, V. Sundaram, G. Elliott, J. Gu, T. Shi, P. Gascard, M. Sigaroudinia, T. D. Tlsty, T. Kadlecek, A. Weiss, H. O’Geen, P. J. Farnham, C. L. Maire, K. L. Ligon, P. A. F. Madden, A. Tam, R. Moore, M. Hirst, M. A. Marra, B. Zhang, J. F. Costello, T. Wang, Functional DNA methylation differences between tissues, cell types, and across individuals discovered using the M&M algorithm. Genome Res. 23, 1522–1540 (2013).

18. W. Wick, M. Platten, C. Meisner, J. Felsberg, G. Tabatabai, M. Simon, G. Nikkhah, K. Papsdorf, J. P. Steinbach, M. Sabel, S. E. Combs, J. Vesper, C. Braun, J. Meixensberger, R. Ketter, R. Mayer-Steinacker, G. Reifenberger, M. Weller, Temozolomide chemotherapy alone versus radiotherapy alone for malignant astrocytoma in the elderly: the NOA-08 randomised, phase 3 trial. Lancet Oncol. 13, 707–715 (2012).

19. K. Ouchi, Y. Sunakawa, A. Tsuji, M. Shiozawa, T. Kawai, H. Yasui, H. Ota, M. Kochi, D. Manaka, H. Ohori, T. Miyake, T. Yamaguchi, M. Matsuura, T. Sagawa, A. Makiyama, M. Takeuchi, W. Ichikawa, M. Fujii, C. Ishioka, Genome-wide DNA methylation status as a biomarker for clinical outcomes of first-line treatment in patients with RAS wild-type metastatic colorectal cancer: JACCRO CC-13AR. ESMO Open 10 (2025).

20. S. Hetzel, A. L. Mattei, H. Kretzmer, C. Qu, X. Chen, Y. Fan, G. Wu, K. G. Roberts, S. Luger, M. Litzow, J. Rowe, E. Paietta, W. Stock, E. R. Mardis, R. K. Wilson, J. R. Downing, C. G. Mullighan, A. Meissner, Acute lymphoblastic leukemia displays a distinct highly methylated genome. Nat. Cancer 3, 768–782 (2022).

21. H. Liao, X. Chen, H. Wang, Y. Lin, L. Chen, K. Yuan, M. Liao, H. Jiang, J. Peng, Z. Wu, J. Huang, J. Li, Y. Zeng, Whole-Genome DNA Methylation Profiling of Intrahepatic Cholangiocarcinoma Reveals Prognostic Subtypes with Distinct Biological Drivers. Cancer Res. 84, 1747–1763 (2024).

22. Y. Zuo, J. Zhong, H. Bai, B. Xu, Z. Wang, W. Li, Y. Chen, S. Jin, S. Wang, X. Wang, R. Wan, J. Xu, K. Fei, J. Han, Z. Yang, H. Bao, Y. Shao, J. Ying, Q. Song, J. Duan, J. Wang, Genomic and epigenomic profiles distinguish pulmonary enteric adenocarcinoma from lung metastatic colorectal cancer. EBioMedicine 82 (2022).

23. N. Gull, M. R. Jones, P. C. Peng, S. G. Coetzee, T. C. Silva, J. T. Plummer, A. L. P. Reyes, B. D. Davis, S. S. Chen, K. Lawrenson, J. Lester, C. Walsh, B. J. Rimel, A. J. Li, I. Cass, Y. Berg, J. P. B. Govindavari, J. K. L. Rutgers, B. P. Berman, B. Y. Karlan, S. A. Gayther, DNA methylation and transcriptomic features are preserved throughout disease recurrence and chemoresistance in high grade serous ovarian cancers. J. Exp. Clin. Cancer Res. 41 (2022).

24. I. Beddows, S. Djirackor, D. K. Omran, E. Jung, N. N. C. Shih, R. Roy, A. Hechmer, A. Olshen, G. Adelmant, A. Tom, J. Morrison, M. Adams, D. C. Rohrer, L. E. Schwartz, C. L. Pearce, H. Auman, J. A. Marto, C. W. Drescher, R. Drapkin, H. Shen, Impact of BRCA mutations, age, surgical indication, and hormone status on the molecular phenotype of the human Fallopian tube. Nature Communications 2025 16:1 16, 1–17 (2025).

25. A. Häkkinen, A. Alkodsi, C. Facciotto, K. Zhang, K. Kaipio, S. Leppä, O. Carpén, S. Grénman, J. Hynninen, S. Hietanen, R. Lehtonen, S. Hautaniemi, Identifying differentially methylated sites in samples with varying tumor purity. Bioinformatics 34, 3078–3085 (2018).

26. O. Kondrashova, M. Topp, K. Nesic, E. Lieschke, G. Y. Ho, M. I. Harrell, G. V. Zapparoli, A. Hadley, R. Holian, E. Boehm, V. Heong, E. Sanij, R. B. Pearson, J. J. Krais, N. Johnson, O. McNally, S. Ananda, K. Alsop, K. J. Hutt, S. H. Kaufmann, K. K. Lin, T. C. Harding, N. Traficante, G. Chenevix-Trench, A. Green, P. Webb, D. Gertig, S. Fereday, S. Moore, J. Hung, K. Harrap, T. Sadkowsky, N. Pandeya, M. Malt, A. Mellon, R. Robertson, T. Vanden Bergh, M. Jones, P. Mackenzie, J. Maidens, K. Nattress, Y. E. Chiew, A. Stenlake, H. Sullivan, B. Alexander, P. Ashover, S. Brown, T. Corrish, L. Green, L. Jackman, K. Ferguson, K. Martin, A. Martyn, B. Ranieri, J. White, V. Jayde, P. Mamers, L. Bowes, L. Galletta, D. Giles, J. Hendley, T. Schmidt, H. Shirley, C. Ball, C. Young, S. Viduka, H. Tran, S. Bilic, L. Glavinas, J. Brooks, R. Stuart-Harris, F. Kirsten, J. Rutovitz, P. Clingan, A. Glasgow, A. Proietto, S. Braye, G. Otton, J. Shannon, T. Bonaventura, J. Stewart, S. Begbie, M. Friedlander, D. Bell, S. Baron-Hay, A. Ferrier, G. Gard, D. Nevell, N. Pavlakis, S. Valmadre, B. Young, C. Camaris, R. Crouch, L. Edwards, N. Hacker, D. Marsden, G. Robertson, P. Beale, J. Beith, J. Carter, C. Dalrymple, R. Houghton, P. Russell, M. Links, J. Grygiel, J. Hill, A. Brand, K. Byth, R. Jaworski, P. Harnett, R. Sharma, G. Wain, B. Ward, D. Papadimos, A. Crandon, M. Cummings, K. Horwood, A. Obermair, L. Perrin, D. Wyld, J. Nicklin, M. Davy, M. K. Oehler, C. Hall, T. Dodd, T. Healy, K. Pittman, D. Henderson, J. Miller, J. Pierdes, P. Blomfield, D. Challis, R. McIntosh, A. Parker, B. Brown, R. Rome, D. Allen, P. Grant, S. Hyde, R. Laurie, M. Robbie, D. Healy, T. Jobling, T. Manolitsas, J. McNealage, P. Rogers, B. Susil, E. Sumithran, I. Simpson, K. Phillips, D. Rischin, S. Fox, D. Johnson, S. Lade, M. Loughrey, N. O’Callaghan, W. Murray, P. Waring, V. Billson, J. Pyman, D. Neesham, M. Quinn, C. Underhill, R. Bell, L. F. Ng, R. Blum, V. Ganju, I. Hammond, Y. Leung, A. McCartney, M. Buck, I. Haviv, D. Purdie, D. Whiteman, N. Zeps, A. deFazio, I. A. McNeish, D. D. Bowtell, E. M. Swisher, A. Dobrovic, M. J. Wakefield, C. L. Scott, Methylation of all BRCA1 copies predicts response to the PARP inhibitor rucaparib in ovarian carcinoma. Nat. Commun. 9 (2018).

27. A. Häkkinen, J. Koiranen, J. Casado, K. Kaipio, O. Lehtonen, E. Petrucci, J. Hynninen, S. Hietanen, O. Carpén, L. Pasquini, M. Biffoni, R. Lehtonen, S. Hautaniemi, qSNE: quadratic rate t-SNE optimizer with automatic parameter tuning for large datasets. Bioinformatics 36, 5086–5092 (2020).

28. K. Lavikka, J. Oikkonen, Y. Li, T. Muranen, G. Micoli, G. Marchi, A. Lahtinen, K. Huhtinen, R. Lehtonen, S. Hietanen, J. Hynninen, A. Virtanen, S. Hautaniemi, Deciphering cancer genomes with GenomeSpy: a grammar-based visualization toolkit. Gigascience 13 (2024).

29. P. A. Jones, Functions of DNA methylation: islands, start sites, gene bodies and beyond. Nat. Rev. Genet. 13, 484–492 (2012).

30. S. Kaluscha, S. Domcke, C. Wirbelauer, M. B. Stadler, S. Durdu, L. Burger, D. Schübeler, Evidence that direct inhibition of transcription factor binding is the prevailing mode of gene and repeat repression by DNA methylation. Nat. Genet. 54, 1895–1906 (2022).

31. H. Guo, J. A. Vuille, B. S. Wittner, E. M. Lachtara, Y. Hou, M. Lin, T. Zhao, A. T. Raman, H. C. Russell, B. A. Reeves, H. M. Pleskow, C. L. Wu, A. Gnirke, A. Meissner, J. A. Efstathiou, R. J. Lee, M. Toner, M. J. Aryee, M. S. Lawrence, D. T. Miyamoto, S. Maheswaran, D. A. Haber, DNA hypomethylation silences anti-tumor immune genes in early prostate cancer and CTCs. Cell 186, 2765–2782.e28 (2023).

32. K. Kitahama, Y. J. Ho, K. Satomi, T. Shibayama, K. Nagahama, K. Ohtsuka, H. Ohnishi, Y. Sakamoto, J. Shibahara, A. Hayashi, Epigenetic evolution and clinicopathological implications of distinct DNA methylation profiles in pancreatic ductal adenocarcinoma. Scientific Reports 2025 15:1 15, 1–13 (2025).

33. F. Blanc-Durand, R. Tang, M. Pommier, M. Nashvi, S. Cotteret, C. Genestie, A. Le Formal, P. Pautier, J. Michels, M. Kfoury, R. Herve, S. Mengue, E. Wafo, A. Elies, G. Miailhe, J. Uzan, E. Rouleau, A. Leary, Clinical Relevance of BRCA1 Promoter Methylation Testing in Patients with Ovarian Cancer. Clin. Cancer Res. 29, 3124–3129 (2023).

34. E. Lengyel, Ovarian cancer development and metastasis. American Journal of Pathology 177, 1053–1064 (2010).

35. I. Konishi, K. Abiko, T. Hayashi, K. Yamanoi, R. Murakami, K. Yamaguchi, J. Hamanishi, T. Baba, N. Matsumura, M. Mandai, Peritoneal dissemination of high-grade serous ovarian cancer: pivotal roles of chromosomal instability and epigenetic dynamics. J. Gynecol. Oncol. 33 (2022).

36. D. W. Garsed, A. Pandey, S. Fereday, C. J. Kennedy, K. Takahashi, K. Alsop, P. T. Hamilton, J. Hendley, Y. E. Chiew, N. Traficante, P. Provan, D. Ariyaratne, G. Au-Yeung, N. W. Bateman, L. Bowes, A. Brand, E. L. Christie, J. M. Cunningham, M. Friedlander, B. Grout, P. Harnett, J. Hung, B. McCauley, O. McNally, A. M. Piskorz, F. A. M. Saner, R. A. Vierkant, C. Wang, S. J. Winham, P. D. P. Pharoah, J. D. Brenton, T. P. Conrads, G. L. Maxwell, S. J. Ramus, C. L. Pearce, M. C. Pike, B. H. Nelson, E. L. Goode, A. DeFazio, D. D. L. Bowtell, The genomic and immune landscape of long-term survivors of high-grade serous ovarian cancer. Nat. Genet. 54, 1853–1864 (2022).

37. S. Y. C. Yang, S. Lheureux, K. Karakasis, J. V. Burnier, J. P. Bruce, D. L. Clouthier, A. Danesh, R. Quevedo, M. Dowar, Y. Hanna, T. Li, L. Lu, W. Xu, B. A. Clarke, P. S. Ohashi, P. A. Shaw, T. J. Pugh, A. M. Oza, Landscape of genomic alterations in high-grade serous ovarian cancer from exceptional long- and short-term survivors. Genome Med. 10 (2018).

38. J. Hopkins, K. Asada, A. Leung, V. Papadaki, H. Davaapil, M. Morrison, T. Orita, R. Sekido, H. Kosuge, M. A. Reddy, K. Kimura, A. Mitani, K. Tsumoto, R. Hamamoto, M. S. Sagoo, S. I. Ohnuma, PRELP Regulates Cell-Cell Adhesion and EMT and Inhibits Retinoblastoma Progression. Cancers (Basel). 14 (2022).

39. H. Schäfer, K. Subbarayan, C. Massa, C. Vaxevanis, A. Mueller, B. Seliger, Correlation of the tumor escape phenotype with loss of PRELP expression in melanoma. J. Transl. Med. 21 (2023).

40. O. Monestier, V. Blanquet, WFIKKN1 and WFIKKN2: “Companion” proteins regulating TGFB activity. Cytokine Growth Factor Rev. 32, 75–84 (2016).

41. D. Afenteva, R. Yu, A. Rajavuori, M. Salvadores, I. M. Launonen, K. Lavikka, K. Zhang, A. Pirttikoski, G. Marchi, S. Jamalzadeh, V. M. Isoviita, Y. Li, G. Micoli, E. P. Erkan, M. M. Falco, D. Ungureanu, A. Lahtinen, J. Oikkonen, S. Hietanen, A. Vähärautio, I. Sur, A. Virtanen, A. Färkkilä, J. Hynninen, T. A. Muranen, J. Taipale, S. Hautaniemi, Multi-omics analysis reveals the attenuation of the interferon pathway as a driver of chemo-refractory ovarian cancer. Cell Rep. Med. 6 (2025).

42. J. A. Ledermann, F. A. Raja, C. Fotopoulou, A. Gonzalez-Martin, N. Colombo, C. Sessa, Newly diagnosed and relapsed epithelial ovarian carcinoma: ESMO clinical practice guidelines for diagnosis, treatment and follow-up. Annals of Oncology 24 (2013).

43. H. Naora, D. J. Montell, Ovarian cancer metastasis: integrating insights from disparate model organisms. Nat. Rev. Cancer 5, 355–366 (2005).

44. D. L. Almeida-Nunes, M. Nunes, H. Osório, V. Ferreira, C. Lobo, P. Monteiro, M. H. Abreu, C. Bartosch, R. Silvestre, R. J. Dinis-Oliveira, S. Ricardo, Ovarian cancer ascites proteomic profile reflects metabolic changes during disease progression. Biochem. Biophys. Rep. 39 (2024).

45. S. Kim, S. Kim, J. Kim, B. Kim, S. I. Kim, M. A. Kim, S. Kwon, Y. S. Song, Evaluating Tumor Evolution via Genomic Profiling of Individual Tumor Spheroids in a Malignant Ascites. Scientific Reports 2018 8:1 8, 1–11 (2018).

46. C. E. Ford, B. Werner, N. F. Hacker, K. Warton, The untapped potential of ascites in ovarian cancer research and treatment. Br. J. Cancer 123, 9–16 (2020).

47. F. Gimeno-Valiente, C. Castignani, E. Larose Cadieux, N. E. Mensah, X. Liu, K. Chen, O. Chervova, T. Karasaki, C. E. Weeden, C. Richard, S. Lai, C. Martínez-Ruiz, E. L. Lim, A. M. Frankell, T. B. K. Watkins, G. Stavrou, I. Usaite, W. T. Lu, D. Marinelli, S. Saghafinia, G. A. Wilson, P. Dhami, H. Vaikkinen, J. Steif, S. Veeriah, R. E. Hynds, M. Hirst, C. Hiley, A. Feber, Ö. Deniz, M. Jamal-Hanjani, N. McGranahan, S. Beck, J. Demeulemeester, M. Tanić, C. Swanton, P. Van Loo, N. Kanu, DNA methylation cooperates with genomic alterations during non-small cell lung cancer evolution. Nature Genetics 2025 57:9 57, 2226–2237 (2025).

48. A. Cervera, V. Rantanen, K. Ovaska, M. Laakso, J. Nuñez-Fontarnau, A. Alkodsi, J. Casado, C. Facciotto, A. Häkkinen, R. Louhimo, S. Karinen, K. Zhang, K. Lavikka, L. Lyly, M. Pal Singh, S. Hautaniemi, Anduril 2: upgraded large-scale data integration framework. Bioinformatics 35, 3815–3817 (2019).

49. A. M. Bolger, M. Lohse, B. Usadel, Trimmomatic: a flexible trimmer for Illumina sequence data. Bioinformatics 30, 2114–2120 (2014).

50. S. Andrews, FastQC. Babraham Institute 0.11.9 [Preprint] (2015).

51. F. Krueger, S. R. Andrews, Bismark: a flexible aligner and methylation caller for Bisulfite-Seq applications. Bioinformatics 27, 1571–1572 (2011).

52. A. R. Quinlan, I. M. Hall, BEDTools: a flexible suite of utilities for comparing genomic features. Bioinformatics 26, 841–842 (2010).

53. A. McKenna, M. Hanna, E. Banks, A. Sivachenko, K. Cibulskis, A. Kernytsky, K. Garimella, D. Altshuler, S. Gabriel, M. Daly, M. A. DePristo, The Genome Analysis Toolkit: a MapReduce framework for analyzing next-generation DNA sequencing data. Genome Res. 20, 1297–1303 (2010).

54. P. Van Loo, S. H. Nordgard, O. C. Lingjærde, H. G. Russnes, I. H. Rye, W. Sun, V. J. Weigman, P. Marynen, A. Zetterberg, B. Naume, C. M. Perou, A. L. Børresen-Dale, V. N. Kristensen, Allele-specific copy number analysis of tumors. Proc. Natl. Acad. Sci. U. S. A. 107, 16910–16915 (2010).

55. F. Perez-Villatoro, J. Oikkonen, J. Casado, A. Chernenko, D. C. Gulhan, M. Tumiati, Y. Li, K. Lavikka, S. Hietanen, J. Hynninen, U. M. Haltia, J. S. Tyrmi, H. Laivuori, P. A. Konstantinopoulos, S. Hautaniemi, L. Kauppi, A. Färkkilä, Optimized detection of homologous recombination deficiency improves the prediction of clinical outcomes in cancer. NPJ Precis. Oncol. 6 (2022).

56. Y. Luo, B. C. Hitz, I. Gabdank, J. A. Hilton, M. S. Kagda, B. Lam, Z. Myers, P. Sud, J. Jou, K. Lin, U. K. Baymuradov, K. Graham, C. Litton, S. R. Miyasato, J. S. Strattan, O. Jolanki, J. W. Lee, F. Y. Tanaka, P. Adenekan, E. O’Neill, J. M. Cherry, New developments on the Encyclopedia of DNA Elements (ENCODE) data portal. Nucleic Acids Res. 48, D882–D889 (2020).

57. P. Danecek, J. K. Bonfield, J. Liddle, J. Marshall, V. Ohan, M. O. Pollard, A. Whitwham, T. Keane, S. A. McCarthy, R. M. Davies, Twelve years of SAMtools and BCFtools. Gigascience 10 (2021).

58. A. Häkkinen, K. Zhang, A. Alkodsi, N. Andersson, E. P. Erkan, J. Dai, K. Kaipio, T. Lamminen, N. Mansuri, K. Huhtinen, A. Vähärautio, O. Carpén, J. Hynninen, S. Hietanen, R. Lehtonen, S. Hautaniemi, PRISM: recovering cell-type-specific expression profiles from individual composite RNA-seq samples. Bioinformatics 37, 2882–2888 (2021).

59. H. M. Finucan, The mode of a multinomial distribution. Biometrika 51, 513–517 (1964).

60. F. Brenet, M. Moh, P. Funk, E. Feierstein, A. J. Viale, N. D. Socci, J. M. Scandura, DNA Methylation of the First Exon Is Tightly Linked to Transcriptional Silencing. PLoS One 6, e14524 (2011).

61. S. S. Wilks, The Large-Sample Distribution of the Likelihood Ratio for Testing Composite Hypotheses. The Annals of Mathematical Statistics 9, 60–62 (1938).

62. L. Van Der Maaten, G. Hinton, Visualizing Data using t-SNE. Journal of Machine Learning Research 9, 2579–2605 (2008).

